# Problems with Evidence Assessment in COVID-19 Health Policy Impact Evaluation: A systematic review of study design and evidence strength

**DOI:** 10.1101/2021.01.21.21250243

**Authors:** Noah A. Haber, Emma Clarke-Deelder, Avi Feller, Emily R. Smith, Joshua Salomon, Benjamin MacCormack-Gelles, Elizabeth M. Stone, Clara Bolster-Foucault, Jamie R. Daw, Laura A. Hatfield, Carrie E. Fry, Christopher B. Boyer, Eli Ben-Michael, Caroline M. Joyce, Beth S. Linas, Ian Schmid, Eric H. Au, Sarah E. Wieten, Brooke A Jarrett, Cathrine Axfors, Van Thu Nguyen, Beth Ann Griffin, Alyssa Bilinski, Elizabeth A. Stuart

**Affiliations:** Meta-Research Innovation Center at Stanford (METRICS), Stanford University, Stanford, CA, USA; Department of Global Health and Population, Harvard T. H. Chan School of Public Health, Boston, MA, USA; Goldman School of Public Policy, UC Berkeley, Berkeley, CA, USA; Department of Global Health, Milken Institute School of Public Health, George Washington University, Washington, D.C, USA; Center for Health Policy and Center for Primary Care and Outcomes Research, Stanford University, Stanford, CA, USA; Department of Health Policy and Management, Johns Hopkins Bloomberg School of Public Health, Baltimore, MD, USA; Epidemiology, Biostatistics, and Occupational Health, McGill University, Montreal, Canada; Health Policy and Management, Columbia University Mailman School of Public Health, New York, NY, USA; Department of Health Care Policy, Harvard Medical School, Boston, MA, USA; Department of Health Policy, Vanderbilt University, Nashville, TN, USA; Department of Epidemiology, Harvard T.H. Chan School of Public Health, Boston, MA, USA; Institute for Quantitative Social Science, Harvard University, Cambridge, MA, USA; Department of Epidemiology, Johns Hopkins Bloomberg School of Public Health, Baltimore, MD, USA; Clinical Quality and Informatics, MITRE Corp, McLean, VA, USA; Department of Mental Health, Johns Hopkins Bloomberg School of Public Health, Baltimore, MD, USA; School of Public Health, University of Sydney, Sydney, Australia; RAND Corporation, Arlington, VA, USA; Interfaculty Initiative in Health Policy, Harvard Graduate School of Arts and Sciences, Cambridge, MA, USA

## Abstract

**Introduction:** Assessing the impact of COVID-19 policy is critical for informing future policies. However, there are concerns about the overall strength of COVID-19 impact evaluation studies given the circumstances for evaluation and concerns about the publication environment. This study systematically reviewed the strength of evidence in the published COVID-19 policy impact evaluation literature.

**Methods:** We included studies that were primarily designed to estimate the quantitative impact of one or more implemented COVID-19 policies on direct SARS-CoV-2 and COVID-19 outcomes. After searching PubMed for peer-reviewed articles published on November 26, 2020 or earlier and screening, all studies were reviewed by three reviewers first independently and then to consensus. The review tool was based on previously developed and released review guidance for COVID-19 policy impact evaluation, assessing what impact evaluation method was used, graphical display of outcomes data, functional form for the outcomes, timing between policy and impact, concurrent changes to the outcomes, and an overall rating.

**Results:** After 102 articles were identified as potentially meeting inclusion criteria, we identified 36 published articles that evaluated the quantitative impact of COVID-19 policies on direct COVID-19 outcomes. The majority (n=23/36) of studies in our sample examined the impact of stay-at-home requirements. Nine studies were set aside because the study design was considered inappropriate for COVID-19 policy impact evaluation (n=8 pre/post; n=1 cross-section), and 27 articles were given a full consensus assessment. 20/27 met criteria for graphical display of data, 5/27 for functional form, 19/27 for timing between policy implementation and impact, and only 3/27 for concurrent changes to the outcomes. Only 1/27 studies passed all of the above checks, and 4/27 were rated as overall appropriate. Including the 9 studies set aside, reviewers found that only four of the 36 identified published and peer-reviewed health policy impact evaluation studies passed a set of key design checks for identifying the causal impact of policies on COVID-19 outcomes.

**Discussion:** The reviewed literature directly evaluating the impact of COVID-19 policies largely failed to meet key design criteria for inference of sufficient rigor to be actionable by policy-makers. This was largely driven by the circumstances under which policies were passed making it difficult to attribute changes in COVID-19 outcomes to particular policies. More reliable evidence review is needed to both identify and produce policy-actionable evidence, alongside the recognition that actionable evidence is often unlikely to be feasible.

## Introduction

Policy decisions to mitigate the impact of COVID-19 on morbidity and mortality are some of the most important issues policymakers have had to make since January 2020. Decisions regarding which policies are enacted depend in part on the evidence base for those policies, including understanding what impact past policies had on COVID-19 outcomes[1,2] Unfortunately, there are substantial concerns that much of the existing literature may be methodologically flawed, which could render its conclusions unreliable for informing policy. The combination of circumstances being difficult for strong impact evaluation, the importance of the topic, and concerns over the publication environment may lead to the proliferation of low strength studies.

High-quality causal evidence requires a combination of rigorous methods, clear reporting, appropriate caveats, and the appropriate circumstances for the methods used.[3–6] Rigorous evidence is difficult in the best of circumstances, and the circumstances for evaluating non-pharmaceutical intervention (NPI) policy effects on COVID-19 are particularly challenging.[5] The global pandemic has yielded a combination of a large number of concurrent policy and non-policy changes, complex infectious disease dynamics, and unclear timing between policy implementation and impact; all of this makes isolating the causal impact of any particular policy or policies exceedingly difficult.[7]

The scientific literature on COVID-19 is exceptionally large and fast growing. Scientists published more than 100,000 papers related to COVID-19 in 2020.[8] There is some general concern that the volume and speed[9,10] at which this work has been produced may result in a literature that is overall low quality and unreliable.[11–15]

Given the importance of the topic, it is critical that decision-makers are able to understand what is known and knowable[5,16] from observational data in COVID-19 policy, as well as what is unknown and/or unknowable.

Motivated by concerns about the methodological strength of COVID-19 policy evaluations, we set out to review the literature using a set of methodological design checks tailored to common policy impact evaluation methods. Our primary objective was to evaluate each paper for methodological strength and reporting, based on pre-existing review guidance developed for this purpose.[17] As a secondary objective, we also studied our own process: examining the consistency, ease of use, and clarity of this review guidance.

This protocol differs in several ways from more traditional systematic review protocols given the atypical objectives and scope of the systematic review. First, this is a systematic review of methodological strength of evidence for a given literature as opposed to a review summary of the evidence of a particular topic. As such, we do not summarize and attempt to combine the results for any of the literature. Second, rather than being a comprehensive review of every possible aspect of what might be considered “quality,” this is a review of targeted critical design features for actionable inference for COVID-19 policy impact evaluation and methods. It is designed to be a set of broad criteria for minimal plausibility of actionable causal inference, where each of the criteria is necessary but not sufficient for strong design. Issues in other domains (data, details of the design, statistics, etc) further reduce overall actionability and quality, and thorough review in those domains is needed for any studies passing our basic minimal criteria. Third, because the scope relies on guided, but difficult and subjective assessments of methodological appropriateness, we utilize a discussion-based consensus process to arrive at consistent and replicable results, rather than a more common model with two independent reviewers with conflict resolution. The independent review serves primarily as a starting point for discussion, but is neither designed nor expected to be a strong indicator of the overall consensus ratings of the group.

## Methods

### Overview

This protocol and study was written and developed following the release of the review guidance written by the author team in September 2020 on which the review tool is based. The protocol for this study was pre-registered on OSF.io[19] in November 2020 following PRISMA guidelines.[20] Deviations from the original protocol are discussed in Appendix 1, and consisted largely of language clarifications and error corrections for both the inclusion criteria and review tool, an increase in the number of reviewers per fully reviewed article from two to three, and simplification of the statistical methods used to assess the data.

For this study, we ascertain minimal criteria for studies to be able to plausibly identify causal effects of policies, which is the information of greatest interest to inform policy decisions. The causal estimand is something that, if known, would definitely help policy makers decide what to do (e.g., whether to implement or discontinue a policy). The study estimates that target causal quantity with a rigorous design and appropriate data in a relevant population/sample. For shorthand, we refer to this as minimal properties of “actionable” evidence.

This systematic review of the strength of evidence took place in three phases: search, screening, and full review.

### Eligibility criteria

The following eligibility criteria were used to determine the papers to include:

- The primary topic of the article must be evaluating one or more individual COVID-19 or *SARS-CoV-2* policies on direct COVID-19 or *SARS-CoV-2* outcomes
  - The primary exposure(s) must be a policy, defined as a government-issued order at any government level to address a directly COVID-19-related outcome (e.g., mask requirements, travel restrictions, etc).
  - *Direct* COVID-19 or *SARS-CoV-2* outcomes *are those that are specific to disease and health outcomes* may include cases detected, mortality, number of tests taken, test positivity rates, Rt, etc.
  - This may NOT include indirect impacts of COVID-19 on items that are not direct *COVID-19 or SARS-CoV-2 impacts* such as income, childcare, *economic impacts, beliefs and attitudes, etc*.
- The primary outcome being examined must be a COVID-19-specific outcome, as above.
- The study must be designed as an impact evaluation study from primary data (i.e., not primarily a predictive or simulation model or meta-analysis).
- The study must be peer reviewed, and published in a peer-reviewed journal indexed by PubMed.
- The study must have the title and abstract available via PubMed at the time of the study start date (November 26).
- The study must be written in English.

These eligibility criteria were designed to identify the literature primarily concerning the quantitative impact of one or more implemented COVID-19 policies on COVID-19 outcomes. Studies in which impact evaluation was secondary to another analysis (such as a hypothetical projection model) were eliminated because they were less relevant to our objectives and/or may not contain sufficient information for evaluation. Categories for types of policies were from the Oxford COVID-19 Government Response Tracker.[21]

### Reviewer recruitment, training, and communication

Reviewers were recruited through personal contacts and postings on online media. All reviewers had experience in systematic review, quantitative causal inference, epidemiology, econometrics, public health, methods evaluation, or policy review. All reviewers participated in two meetings in which the procedures and the review tool were demonstrated. Screening reviewers participated in an additional meeting specific to the screening process. Throughout the main review process, reviewers communicated with the administrators and each other through Slack for any additional clarifications, questions, corrections, and procedures. The main administrator (NH), who was also a reviewer, was available to answer general questions and make clarifications, but did not answer questions specific to any given article.

### Review phases and procedures

#### Search strategy

The search terms combined four Boolean-based search terms: a) COVID-19 research,17 b) regional government units (e.g., country, state, county, and specific country, state, or province, etc.), c) policy or policies, and d) impact or effect. The full search terms are available in Appendix 2.

#### Information Sources

The search was limited to published articles in peer-reviewed journals. This was largely to attempt to identify literature that was high quality, relevant, prominent, and most applicable to the review guidance. PubMed was chosen as the exclusive indexing source due to the prevalence and prominence of policy impact studies in the health and medical field. Preprints were excluded to limit the volume of studies to be screened and to ensure each had met the standards for publication through peer review. The search was conducted on November 26, 2020.

#### Study Selection

Two reviewers were randomly selected to screen the title and abstract of each article for the inclusion criteria. In the case of a dispute, a third randomly selected reviewer decided on acceptance/rejection. Eight reviewers participated in the screening. Training consisted of a one-hour instruction meeting, a review of the first 50 items on each reviewers’ list of assigned articles, and a brief asynchronous online discussion before conducting the full review.

#### Full article review

The full article review consisted of two sub-phases: the independent primary review phase, and a group consensus phase. The independent review phase was designed primarily for the purpose of supporting and facilitating discussion in the consensus discussion, rather than as high stakes definitive review data on its own. The consensus process was considered the primary way in which review data would be generated, rather than synthesis from the independent reviews. A flow diagram of the review process is available in Appendix 3

Each article was randomly assigned to three of the 23 reviewers in our review pool. Each reviewer independently reviewed each article on their list, first for whether the study met the eligibility criteria, then responding to methods identification and guided strength of evidence questions using the review tool, as described below. Reviewers were able to recuse themselves for any reason, in which case another reviewer was randomly selected. Once all three reviewers had reviewed a given article, all articles that weren’t unanimously determined to not meet the inclusion criteria underwent a consensus process.

During the consensus round, the three reviewers were given all three primary reviews for reference, and were tasked with generating a consensus opinion among the group. One randomly selected reviewer was tasked to act as the arbitrator. The arbitrator’s primary task was facilitating discussion and for moving the group toward establishing a consensus that represented the collective subjective assessments of the group. If consensus could not be reached, a fourth randomly selected reviewer was brought into the discussion to help resolve disputes.

### Review tool for data collection

This review tool and data collection process was an operationalized and lightly adapted version of the COVID-19 health policy impact evaluation review guidance literature, written by the lead authors of this study. The main adaptation was removing references to the COVID-19 literature. All reviewers were instructed to read and refer to this guidance document to guide their assessments. The full guidance manuscript contains additional explanation and rationale for all parts of this review and the tool, and is available both in the adapted form as was provided to the reviewers in a supplementary file “CHSPER review guidance refs removed.pdf” and in an updated version in Haber et al., 2020.[17] The full review tool is attached as supplementary file “review tool final.pdf”.

The review tool consisted of two main parts: methods design categorization and full review. The review tool and guidance categorizes policy causal inference designs based on the structure of their assumed counterfactual. This is assessed through identifying the data structure and comparison(s) being made. There are two main items for this determination: the number of pre-period time points (if any) used to assess pre-policy outcome trends, and whether or not policy regions were compared with non-policy regions. These, and other supporting questions, broadly allowed categorization of methods into cross-sectional, pre/post, interrupted time series (ITS), difference-in-differences (DiD), comparative interrupted time-series (CITS), (randomized) trials, or other. Given that most papers have several analyses, reviewers were asked to focus exclusively on the impact evaluation analysis that was used as the primary support for the main conclusion of the article.

Studies categorized as cross-sectional, pre/post, randomized controlled trial designs, and other were included in our sample, but set aside for no further review for the purposes of this research. Cross-sectional and pre/post studies are not considered sufficient to yield well-identified causal inference in the specific context of COVID-19 policy impact evaluation, as explained in the policy impact evaluation guidance documentation. Cross-sectional and pre-post designs were considered inappropriate for policy causal inference for COVID-19 due largely to inability to account for a large number of potential issues, including confounding, epidemic trends, and selection biases. Randomized controlled trials were assumed to broadly meet key design checks. Studies categorized as “other” received no further review, as the review guidance would be unable to assess them. Additional justification and explanation for this decision is available in the review guidance.

For the methods receiving full review (ITS, DiD, and CITS), reviewers were asked to identify potential issues and give a category-specific rating. The specific study designs triggered sub-questions and/or slightly altered the language of the questions being asked, but all three of the methods design categories shared these four key questions:

- Graphical presentation: “Does the analysis provide graphical representation of the outcome over time?”
  - Graphical presentation refers to how the authors present the data underlying their impact evaluation method. This is a critical criteria for assessing the potential validity of the assumed model. The key questions here are whether any chart shows the outcome over time and the assumed models of the counterfactuals. To meet a high degree of confidence in this category, graphical displays must show the outcome and connect to the counterfactual construction method.
- Functional form: “Is the functional form of the model used for the trend in counterfactual infectious disease outcomes (e.g., linear, non-parametric, exponential, logarithmic, etc.,) well-justified and appropriate?”
  - Functional form refers to the statistical functional form of the trend in counterfactual infectious disease outcomes (i.e. the assumptions used to construct counterfactual outcomes). This may be a linear function, non-parametric, exponential or logarithmic function, infectious disease model projection, or any other functional form. The key criteria here are whether this is discussed and justified in the manuscript, and if so, is it a plausibly appropriate choice given infectious disease outcomes.
- Timing of policy impact: “Is the date or time threshold set to the appropriate date or time (e.g., is there lag between the intervention and outcome)?”
  - Timing of policy impact refers to assumptions about when we would expect to see an impact from the policy vis-a-vis the timing of the policy introduction. This would typically be modelled with leads and lags. The impact of policy can occur before enactment (e.g., in cases where behavior change after policy is announced, but before it takes place in anticipation) or long after the policy is enacted (e.g., in cases where it takes time to ramp up policy implementation or impacts). The key criteria here are whether this is discussed and justified in the manuscript, and if so, whether it is a plausibly appropriate choice given the policy and outcome.
- Concurrent changes: “Is this policy the only uncontrolled or unadjusted-for way in which the outcome could have changed during the measurement period [differently for policy and non-policy regions]?”
  - Concurrent changes refers to the presence of uncontrolled other events and changes that may influence outcomes at the same time as the policy would impact outcomes. In order to assess the impact of one policy or set of policies, the impact of all other forces that differentially impact the outcome must either be negligible or controlled for. The key criteria here are whether it is likely that there are substantial other uncontrolled forces (e.g. policies, behavioral changes, etc) which may be differentially impacting outcomes at the same time as the policy of interest.

For each of the four key questions, reviewers were given the option to select “No,” “Mostly no,” “Mostly yes,” and “Yes” with justification text requested for all answers other than “Yes.” Each question had additional prompts as guidance, and with much more detail provided in the full guidance document. Ratings are, by design, subjective assessments of the category according to the guidance. We do not use numerical scoring, for similar reasons as Cochrane suggests that the algorithms for summary judgements for the RoB2 tool are merely “proposed” assessments, which reviewers should change as they believe appropriate.[22] It is entirely plausible, for example, for a study to meet all but one criteria but for the one remaining to be sufficiently violated that the entire collective category is compromised. Alternatively, there could be many minor violations of all of the criteria, but that they were collectively not sufficiently problematic to impact overall ratings. Further, reviewers were also tasked with considering room for doubt in cases where answers to these questions were unclear.

The criteria were designed to establish minimal plausibility of actionable evidence, rather than certification of high quality. Graphical representation is included here primarily as a key way to assess the plausibility and justification of key model assumptions, rather than being necessary for validity by itself. For example, rather than having the “right” functional form or lag structure, the review guidance asks whether the functional form and lags is discussed at all and (if discussed) reasonable.

These four questions were selected and designed being critical to evaluating strength of study design for policy impact evaluation in general, direct relevance for COVID-19 policy, feasibility for use in guided review. These questions are designed as minimal and key criteria for plausibly actionable impact evaluation design for COVID-19 policy impact evaluation, rather than as a comprehensive tool assessing overall quality. Thorough review of data quality, statistical validity, and other issues are also critical points of potential weakness in study designs, and would be needed in addition to these criteria, if these key design criteria are met. A thorough justification and explanation of how and why these questions were selected is available in the provided guidance document and in Haber et al., 2020.[17]

Finally, reviewers were asked a summary question:

- Overall: “Do you believe that the design is appropriate for identifying the policy impact(s) of interest?”

Reviewers were asked to consider the scale of this question to be both independent/not relative to any other papers, and that any one substantial issue with the study design could render it a “No” or “Mostly no.” Reviewers were asked to follow the guidance and their previous answers, allowing for their own weighting of how important each issue was to the final result. A study could be excellent on all dimensions except for one, and that one dimension could render it inappropriate for causal inference. As such, in addition to the overall rating question, we also generated a “weakest link” metric for overall assessment, representing the lowest rating among the four key questions (graphical representation, functional form, timing of policy impact, and concurrent changes). A “mostly yes” or “yes” is considered a passing rating, indicating that the study was not found to be inappropriate on the specific dimension of interest.

A “yes” rating does not necessarily indicate that the study is strongly designed, conducted, or is actionable; it only means that it passes a series of key design checks for policy impact evaluation and should be considered for further evaluation. The papers may contain any number of other issues that were not reviewed (e.g., statistical issues, inappropriate comparisons, generalizability, etc.,). As such, this should only be considered an initial assessment of plausibility that the study is well-designed, rather than confirmation that it is appropriate and applicable.

The full review tool is available in the supplementary materials.

#### Heterogeneity

Inter-rater reliability (IRR) was assessed using Krippendorff’s alpha.[23,24] Rather than more typical uses intended as an examination of the “validity” of ratings, the IRR statistic in this case is being used as a heuristic indicator of heterogeneity between reviewers during the independent phase, where heterogeneity is both expected and not necessarily undesirable. As a second examination of reviewer heterogeneity, we also show the distribution of category differences between primary reviewers within a study (e.g. if primary reviewers rated “Yes,” “Mostly no,” and “Mostly yes” there are two pairs of answers that were one category different, and one pair that was two categories different).

### Statistical analysis

Statistics provided are nearly exclusively counts and percentages of the final dataset. Analyses and graphics were performed in R.[25] Krippendorff’s alpha was calculated using the IRR package.[26] Relative risks were estimated using the epitools package.[27]

Citation counts for accepted articles were obtained through Google Scholar[28] on January 11, 2021. Journal impact factors were obtained from the 2019 Journal Citation Reports.[29]

### Data and code

Data, code, the review tool, and the review guidance are stored and available here: https://osf.io/9xmke/files/. The dataset includes full results from the search and screening and all review tool responses from reviewers during the full review phase.

## Results

### Search and screening

After search and screening of titles and abstracts, 102 articles were identified as likely or potentially meeting our inclusion criteria. Of those 102 articles, 36 studies met inclusion after independent review and deliberation in the consensus process. The most common reasons for rejection at this stage were that the study did not measure the quantitative direct impact of specific policies and/or that such an impact was not the main purpose of the study. Many of these studies implied that they measured policy impact in the abstract or introduction, but instead measured correlations with secondary outcomes (e.g., the effect of movement reductions, which are influenced by policy) and/or performed cursory policy impact evaluation secondary to projection modelling efforts.

### Descriptive statistics

Publication information from our sample is shown in Figure 2. The articles in our sample were generally published in journals with high impact factors (median impact factor: 3.6, 25th percentile: 2.3, 75th percentile: 5.3 IQR: 3.0) and have already been cited in the academic literature (median citation count: 5.0, 25th percentile: 2.0, 75th percentile: 26.8, IQR 24.8, on 1/11/21). The most commonly evaluated policy type was stay at home requirements (64% n=23/36). Reviewers noted that many articles referenced “lockdowns,” but did not define the specific policies to which this referred. Reviewers specified mask mandates for 3 of the studies, and noted either a combination of many interventions or unspecified specific policies in 7 cases.

**Figure 1:**
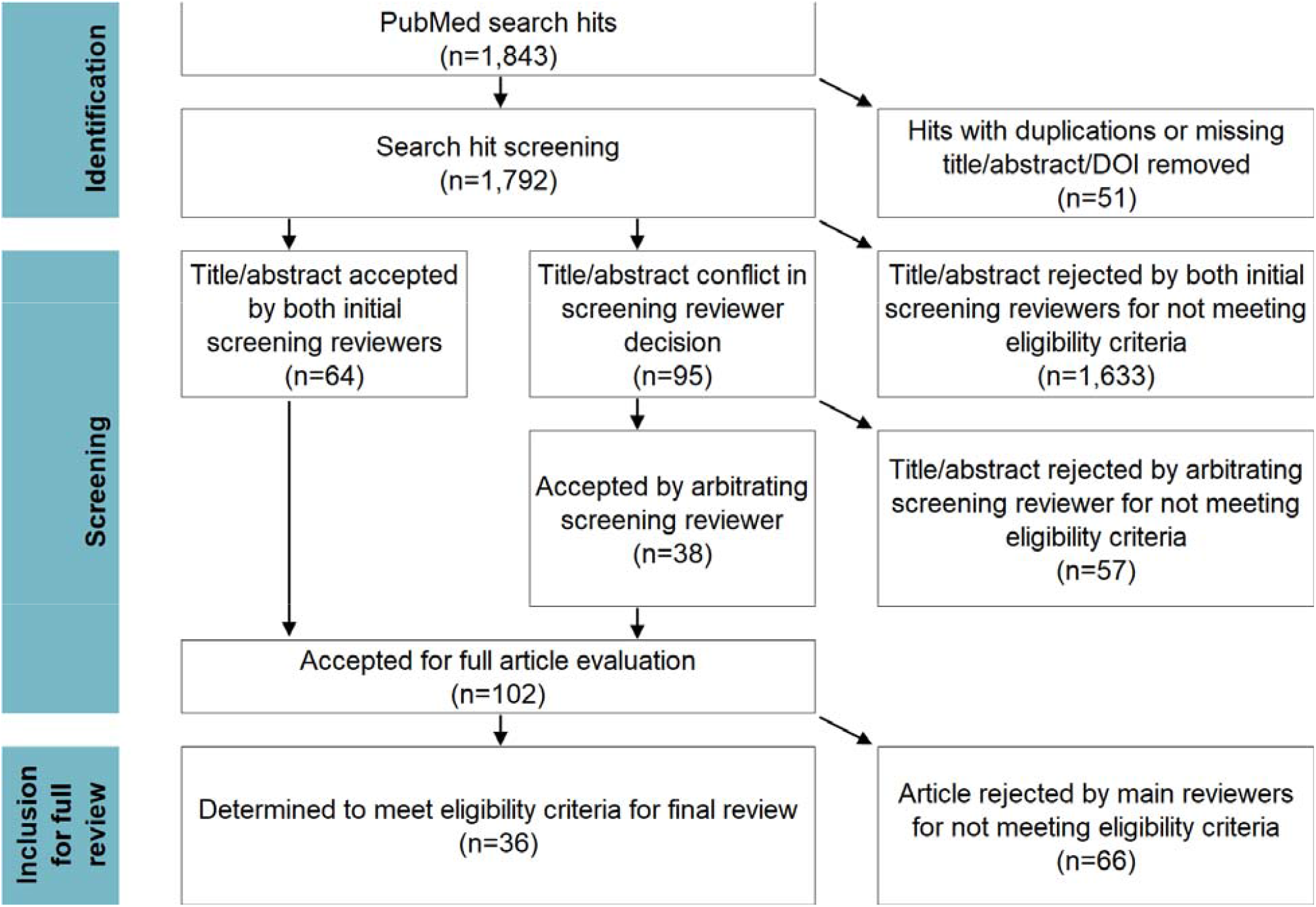
PRISMA diagram of systematic review process.

**Figure 2:**
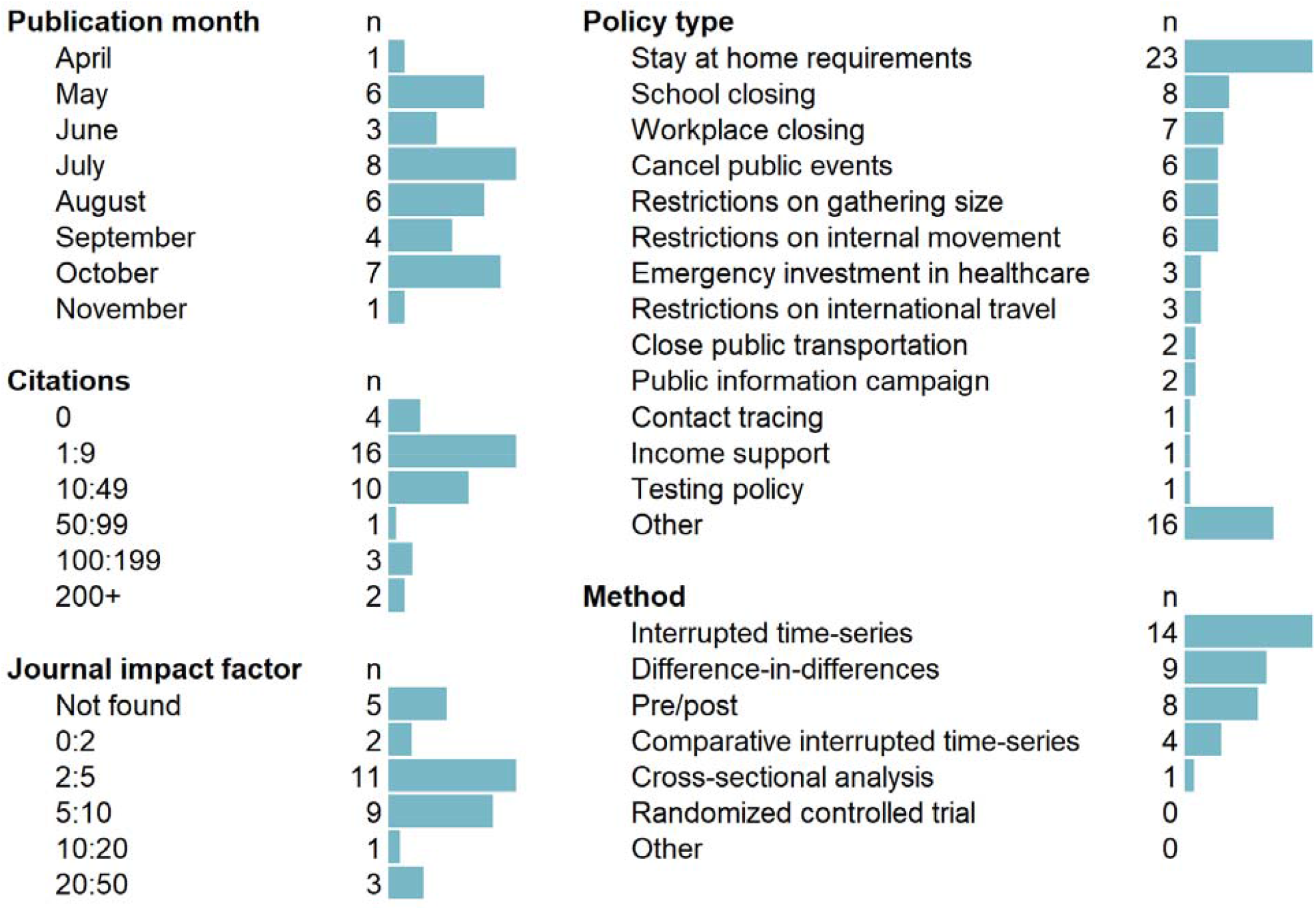
Descriptive sample statistics (n=36)

Reviewers most commonly selected interrupted time-series (39% n=14/36) as the methods design, followed by difference-in-differences (9% n=9/36) and pre-post (8% n=8/36). There were no randomized controlled trials of COVID-19 health policies identified (0% n=0/36), nor were any studies identified that reviewers could not categorize based on the review guidance (0% n=0/36).

The identified articles and selected review results are summarized in Table 1.

**Table 1:**
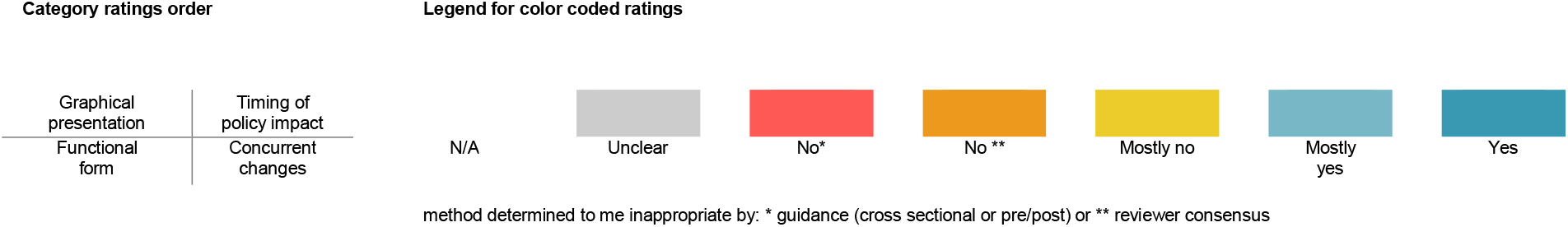
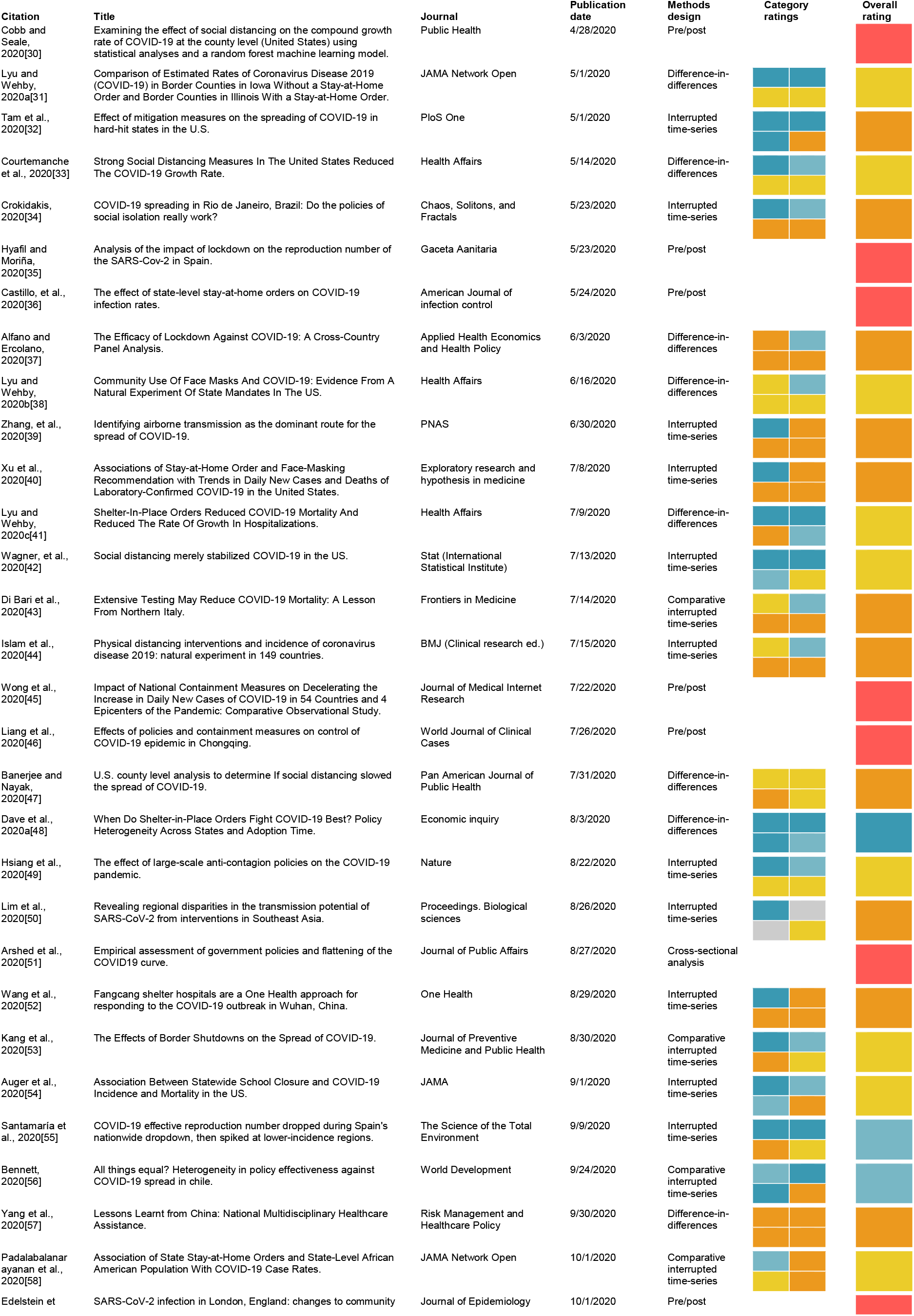
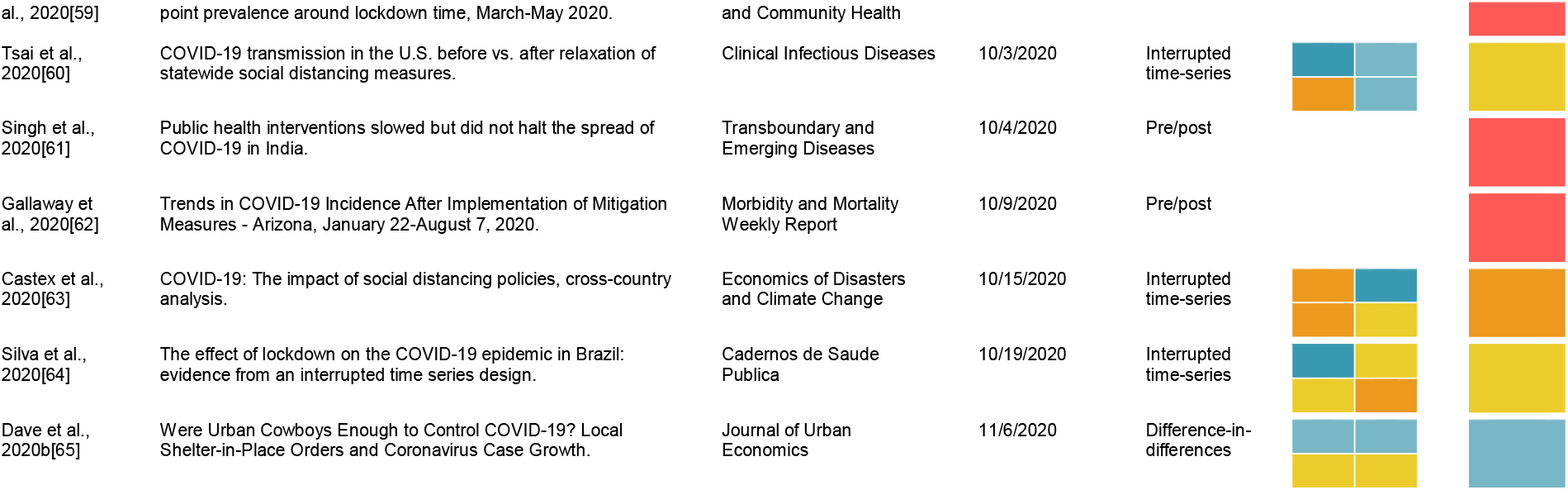
Summary of articles reviewed and reviewer ratings for key and overall questions

### Strength of methods assessment

Graphical representation of the outcome over time was relatively well-rated in our sample, with 74% (n=20/27) studies being given a “mostly yes” or “yes” rating for appropriateness. Reasons cited for non-”yes” ratings included a lack of graphical representation of the data, alternative scales used, and not showing the dates of policy implementation.

Functional form issues appear to have presented a major issue in these studies, with only 19% receiving a “mostly yes” or “yes” rating, 78% (n=21/27) receiving a “no” rating, and 4% (n=1/27) “unclear.” There were two common themes in this category: studies generally using scales that were broadly considered inappropriate for infectious disease outcomes (e.g., linear counts), and/or studies lacking stated justification for the scale used. Reviewers also noted disconnects between clear curvature in the outcomes in the graphical representations and the analysis models and outcome scales used (e.g., linear). In one case, reviewers could not identify the functional form actually used in analysis.

Reviewers broadly found that these studies dealt with timing of policy impact (e.g., lags between policy implementation and expected impact) relatively well, with 70% (n=19/27) rated “yes” or “mostly yes.” Reasons for non-”yes” responses included not adjusting for lags and a lack of justification for the specific lags used.

Concurrent changes were found to be a major issue in these studies, with only 11% (n=3/27) studies receiving passing ratings (“yes” or “mostly yes”) with regard to uncontrolled concurrent changes to the outcomes. Reviewers nearly ubiquitously noted that the articles failed to account for the impact of other policies that could have impacted COVID-19 outcomes concurrent with the policies of interest. Other issues cited were largely related to non-policy-induced behavioral and societal changes.

When reviewers were asked if sensitivity analyses had been performed on key assumptions and parameters, about half (56% n=15/27) answered “mostly yes” or “yes.” The most common reason for non-”yes” ratings was that, while sensitivity analyses were performed, they did not address the most substantial assumptions and issues.

Overall, reviewers rated only four studies (11%, n=4/36,) as being plausibly appropriate (“mostly yes” or “yes”) for identifying the impact of specific policies on COVID-19 outcomes, as shown in Figure 3. 25% (n=9/36) were automatically categorized as being inappropriate due to being either cross-sectional or pre/post in design, 33% (n=12/36) of studies were given a “no” rating for appropriateness, 31% “mostly no” (n=11/36), 8% “mostly yes” (n=3/36), and 3% “yes” (n=1/36). The most common reason cited for non-”yes” overall ratings was failure to account for concurrent changes (particularly policy and societal changes).

**Figure 3:**
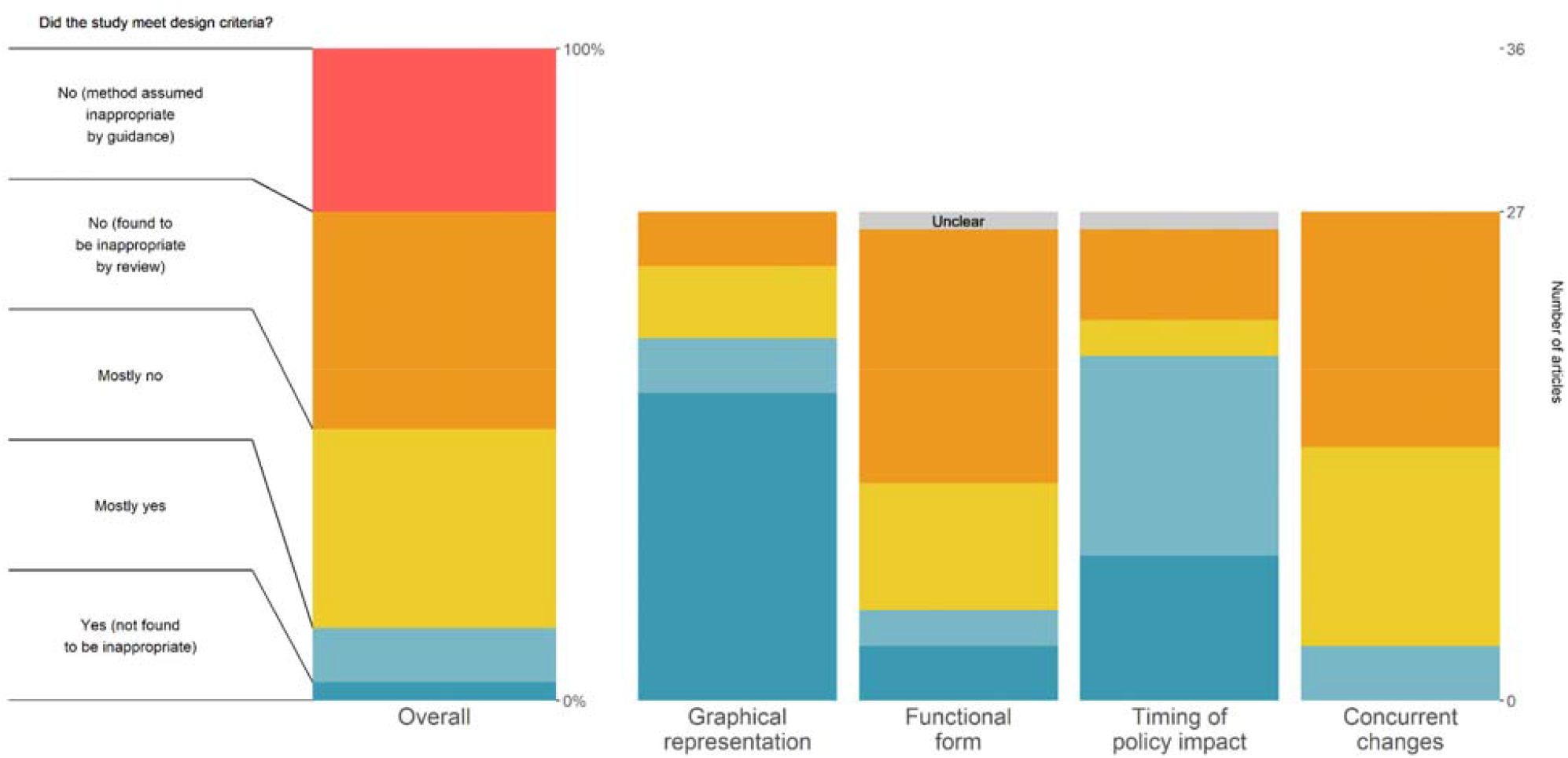
Main consensus results summary for key and overall questions. This chart shows the final overall ratings (left) and the key design question ratings for the consensus review of the 36 included studies, answering the degree to which the articles met the given key design question criteria. The key design question ratings were not asked for the nine included articles which selected methods assumed by the guidance to be non-appropriate. The question prompt in the figure is shortened for clarity, where the full prompt for each key question is available in the Methods section.

As shown in Figure 4, the consensus overall proportion passing (“mostly yes” or “yes”) was a quarter of what it was from the initial independent reviews. 45% (n=34/75) of studies were rated as “yes” or “mostly yes” in the initial independent review, as compared to 11% (n=4/36) in the consensus round (RR 0.25, 95%CI 0.09:0.64). The issues identified and discussed in combination during consensus discussions, as well as additional clarity on the review process, resulted in reduced overall confidence in the findings. Increased clarity on the review guidance with experience and time may also have reduced these ratings further.

**Figure 4:**
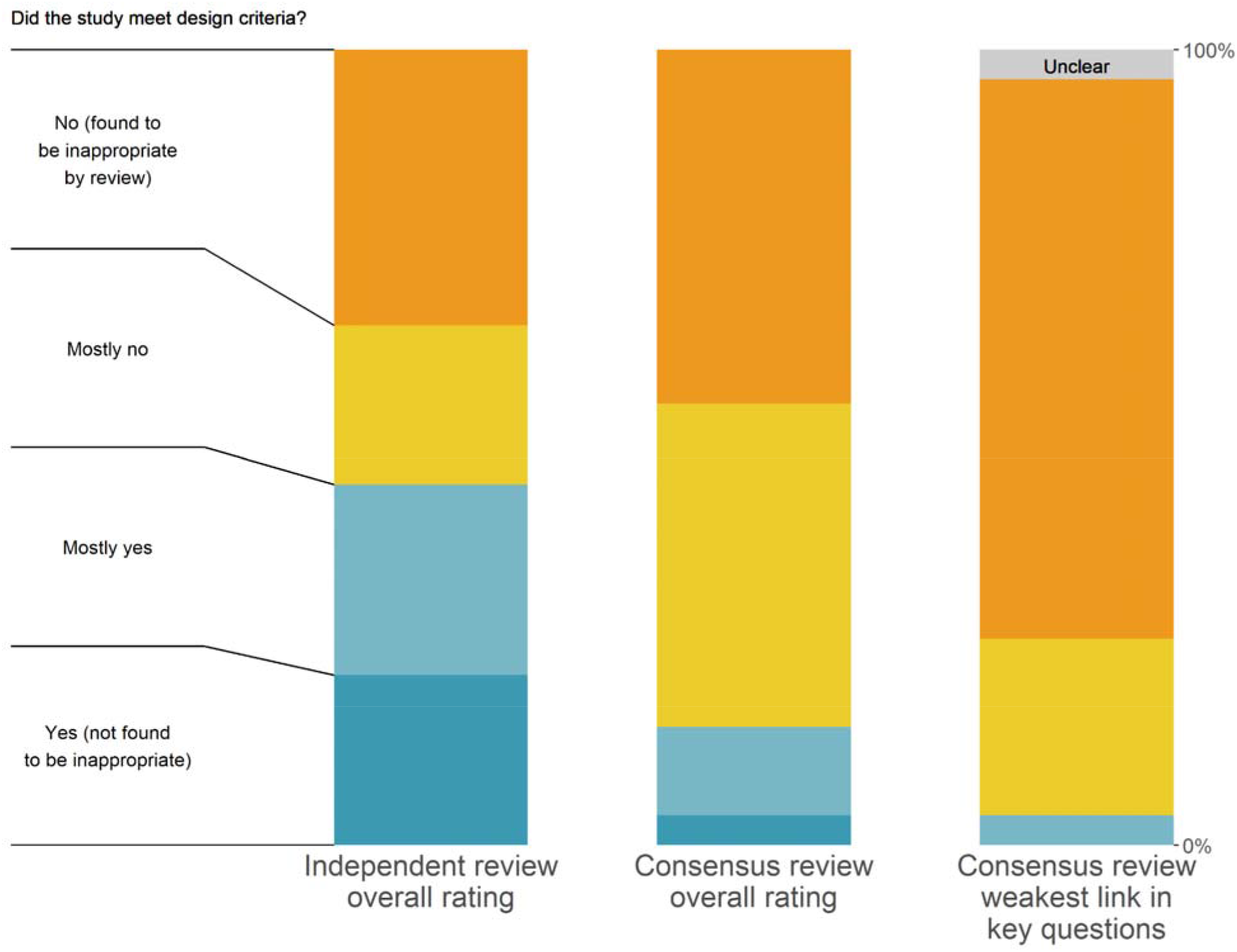
Comparison of independent reviews, weakest link, and direct consensus review. This chart shows the final overall ratings by three different possible metrics. The first column contains all of the independent review ratings for the 27 studies which were eventually included in our sample, noting that reviewers who either selected them as not meeting inclusion criteria or selected a method that didn’t receive the full review did not contribute. The middle column contains the final consensus reviews among the 27 articles which received full review. The last column contains the weakest link rating, as described in the methods section. The question prompt in the figure is shortened for clarity, where the full prompt for each key question is available in the Methods section.

The large majority of studies had at least one “no” or “unclear” rating in one of the four categories (74% n=20/27), with only one study whose lowest rating was a “mostly yes,” no studies rated “yes” in all four categories. Only one study was found to pass design criteria in all four key questions categories, as shown in the “weakest link” column in Figure 4.

### Review process assessment

During independent review, all three reviewers independently came to the same conclusions on the main methods design category for 33% (n=12/36) articles, two out of the three reviewers agreed for 44% (n=16/36) articles, and none of the reviewers agreed in 22% (n=8/36) cases. One major contributor to these discrepancies were the 31% (n=11/36) cases where one or more reviewers marked the study as not meeting eligibility criteria, 64% (n=7/11) of which the other two reviewers agreed on the methods design category.

Reviewers’ initial independent reviews were heterogeneous for key rating questions. For the overall scores, Krippendorff’s alpha was only 0.16 due to widely varying opinions between raters. The four key categorical questions had slightly better inter-rater reliability than the overall question, with Krippendoff’s alphas of 0.59 for graphical representation, 0.34 for functional form, 0.44 for timing of policy impact, and 0.15 for concurrent changes, respectively.For the main summary rating, primary reviewers within each study agreed in 26% of cases (n=16), were one category different in 45% (n=46), two categories different in 19% (n=12), and three categories (i.e. the maximum distance, “Yes” vs “No”) in 10% of cases (n=6).

The consensus rating for overall strength was equal to the lowest rating among the independent reviews in 78% (n=21/27) of cases, and only one higher than the lowest in the remaining 22% (n=6/27). This strongly suggests that the multiple reviewer review, discussion, and consensus process more thoroughly identifies issues than independent review alone. There were two cases for which reviewers requested an additional fourth reviewer to help resolve standing issues for which the reviewers felt they were unable to come to consensus.

The most consistent point of feedback from reviewers was the value of having a three reviewer team with whom to discuss and deliberate, rather than two as initially planned. This was reported to help catch a larger number of issues and clarify both the papers and the interpretation of the review tool questions. Reviewers also expressed that one of the most difficult parts of this process was assessing the inclusion criteria, some of the implications of which are discussed below.

## Discussion

This systematic review of evidence strength found that only four (or only one by a stricter standard) of the 36 identified published and peer-reviewed health policy impact evaluation studies passed a set of key checks for identifying the causal impact of policies on COVID-19 outcomes. Because this systematic review examined a limited set of key study design features and did not address more detailed aspects of study design, statistical issues, generalizability, and any number of other issues, this result may be considered an upper bound on the overall strength of evidence within this sample. Two major problems are nearly ubiquitous throughout this literature: failure to isolate the impact of the policy(s) of interest from other changes that were occurring contemporaneously, and failure to appropriately address the functional form of infectious disease outcomes in a population setting. While policy decisions are being made on the backs of high impact-factor papers, we find that the citation-based metrics do not correspond to “quality” research as used by Yin et al., 2021.[66] Similar to other areas in the COVID-19 literature,[67] we found the current literature directly evaluating the impact of COVID-19 policies largely fails to meet key design criteria for actionable inference to inform policy decisions.

The framework for the review tool is based on the requirements and assumptions built into policy evaluation methods. Quasi-experimental methods rely critically on the scenarios in which the data are generated. These assumptions and the circumstances in which they are plausible are well-documented and understood,[2,4–6,18,68] including one paper discussing application of difference-in-differences methods specifically for COVID-19 health policy, released in May 2020.[5] While “no uncontrolled concurrent changes” is a difficult bar to clear, that bar is fundamental to inference using these methods.

The circumstances of isolating the impact of policies in COVID-19 - including large numbers of policies, infectious disease dynamics, and massive changes to social behaviors - make those already difficult fundamental assumptions broadly much less likely to be met. Some of the studies in our sample were nearly the best feasible studies that could be done given the circumstances, but the best that can be done often yields little actionable inference. The relative paucity of strong studies does not in any way imply a lack of impact of those policies; only that we lack the circumstances to have evaluated their effects.

Because the studies estimating the harms of policies share the same fundamental circumstances, the evidence of COVID-19 policy harms is likely to be of similarly poor strength. Identifying the effects of many of these policies, particularly for the spring of 2020, is likely to be unknown and perhaps unknowable. However, there remains additional opportunities with more favorable circumstances, such as measuring overall impact of NPIs as bundles, rather than individual policies. Similarly, studies estimating the impact of re-opening policies or policy cancellation are likely to have fewer concurrent changes to address.

The review process itself demonstrates how guided and targeted peer review can efficiently evaluate studies in ways that the traditional peer review systems do not. The studies in our sample had passed the full peer review process, were published in largely high-profile journals, and are highly cited, but contained substantial flaws that rendered their inference utility questionable. The relatively small number of studies included, as compared to the size of the literature concerning itself with COVID-19 policy, may suggest that there was relative restraint from journal editors and reviewers for publishing these types of studies. The large number of models, but relatively small number of primary evaluation analyses is consistent with other areas of COVID-19.[69,70] At minimum, the flaws and limitations in their inference could have been communicated at the time of publication, when they are needed most. In other cases, it is plausible that many of these studies would not have been published had a more thorough or more targeted methodological review been performed.

This systematic review of evidence strength has limitations. The tool itself was limited to a very narrow - albeit critical - set of items. Low ratings in our study should not be interpreted as being overall poor studies, as they may make other contributions to the literature that we did not evaluate. While the guidance and tool provided a well-structured framework and our reviewer pool was well-qualified, strength of evidence review is inherently subjective. It is plausible and likely that other sets of reviewers would come to different conclusions for each study, but unlikely that the overall conclusions of our assessment would change substantially. However, the consensus process was designed with these issues subjectivity in mind, and demonstrates the value of consensus processes for overcoming hurdles with subjective and difficult decisions.

While subjective assessments are inherently subject to the technical expertise, experiences, and opinions of reviewers, we argue they are both appropriate and necessary to reliably assess strength of evidence based on theoretical methodological issues. With the exception of the graphical assessment, proper assessment of the core methodological issues requires that reviewers are able to weigh the evidence as they see fit. Much like standard institutional peer review, reviewers independently had highly heterogeneous opinions, attributable to differences in opinion or training, misunderstandings/learning about the review tool and process, and expected reliance on the consensus process. Unlike traditional peer review, there was subject-matter-specific guidance and a process to consolidate and discuss those heterogenous initial opinions. The reduction in ratings from the initial highly heterogeneous ratings to a lower heterogeneity in ratings indicates that reviewers had initially identified issues differently, but that the discussion and consensus process helped elucidate the extent of the different issues that each reviewer detected and brought to discussion. This also reflects reviewer learning over time, where reviewers were better able to identify issues at the consensus phase than earlier. It is plausible that stronger opinions had more weight, but we expect that this was largely mitigated by the random assignment of the arbitrator, and reviewer experiences did not indicate this as an issue.

Most importantly, this review does not cover all policy inference in the scientific literature. One large literature from which there may be COVID-19 policy evaluation otherwise meeting our inclusion criteria are pre-prints. Many pre-prints would likely fare well in our review process. Higher strength papers often require more time for review and publication, and many high quality papers may be in the publication pipeline now. Second, this review excluded studies that had a quantitative impact evaluation as a secondary part of the study (e.g., to estimate parameters for microsimulation or disease modeling). Third, the review does not include policy inference studies that do not measure the impact of a specific policy. For instance, there are studies that estimate the impact of reduced mobility on COVID-19 outcomes but do not attribute the reduced mobility to any specific policy change. Finally, a considerable number of studies that present analyses of COVID-19 outcomes to inform policy are excluded because they do not present a quantitative estimate of specific policies’ treatment effects.

While COVID-19 policy is one of the most important problems of our time, the circumstances under which those policies were enacted severely hamper our ability to study and understand their effects. Claimed conclusions are only as valuable as the methods by which they are produced. Replicable, rigorous, intense, and methodologically guided review is needed to both communicate our limitations and make more actionable inference. Weak, unreliable, and overconfident evidence leads to poor decisions and undermines trust in science.[15,71] In the case of COVID-19 health policy, a frank appraisal of the strength of the studies on which policies are based is needed, alongside the understanding that we often must make decisions when strong evidence is not feasible.[72]

## Data Availability

Data, code, the review tool, and the review guidance are stored and available here: https://osf.io/9xmke/files/

## Acknowledgements

We would like to thank Dr. Steven Goodman and Dr. John Ioannidis for their support during the development of this study, and Dr. Lars Hemkins and Dr. Mario Malicki for helpful comments in the protocol development.

## Author roles

Screening reviewers: CJ, SW, CB, CA, NH, CBF, VN, and Keletso Makofane

Full article reviewers: NH, ECD, AF, BMG, ES, CBF, JD, LH, CG, CB, EBM, CJ, BL, IS, EA, SW, BJ, CA, VN, BG, AB, ES

Protocol development: NH, EC, JS, AF, ES

Administration, primary manuscript writing, data management, and analysis: NH

All authors participated in manuscript editing.

## Appendix 1: Changes from pre-registered protocol and justifications

The full, original pre-registered protocol is available here: https://osf.io/7nbk6

### Inclusion criteria

Minor language edits were made to the inclusion criteria to improve clarity and fix grammatical and typographical errors. This largely centered around improving clarity that a study must estimate the quantitative impact of policies that had already been enacted. The word “quantitative” was not explicitly stated in the original version.

### Procedures

The original protocol specified that each article would receive two independent reviewers. This was increased to three reviewers per article once it became clear both that the number of articles which would be accepted for full review was lower than expectations, and that there would be substantial differences in opinion between reviewers.

### Statistical analysis

Firstly, the original protocol specified that 95% confidence intervals would be calculated. However, after further discussion and review, we determined that sampling-based confidence intervals were not appropriate. Our results are not indicative nor intended to be representative of any super- or target-population, and as such sampling-based error is not an appropriate metric for the conclusions of this study.

Secondly, the original protocol specified Kappa-based interrater reliability statistics. However, using three reviewers, rather than the originally registered two, meant that most Kappa statistics would not be appropriate for our review process. Given the three-rater, four-level ordinal scale used, we opted instead to use Krippendorff’s Alpha.

### Review tool

A number of changes were made to the review tool during the course of the review process. While the original protocol included logic to allow pre/post for review in some of the key questions, this was removed for consistency with the guidance document.

The remaining changes to the review tool were error corrections and clarifications (e.g. correcting the text for the concurrent changes sections in difference-in-differences so that it stated “uncontrolled” concurrent changes, and distinguishing the DiD/CITS requirements from the ITS requirements to emphasize differential concurrent changes).

## Appendix 2: Full search terms

Note: The search filter for COVID-19 and SARS-CoV-2 were the exact search terms used for the National Library of Medicine one-click search option at the time of the protocol development and when the search took place. This reflects that some of the early literature referred to Wuhan specifically (both in geographic reference for where the SARS-CoV-2 was initially found, and unfortunately also early naming of the virus/disease) before official naming conventions became ubiquitous in the literature. In order to comprehensively capture the literature and use searching best practices, we used the most standard and recommended terms.

((((wuhan[All Fields] AND (“coronavirus”[MeSH Terms] OR “coronavirus”[All Fields])) AND 2019/12[PDAT] : 2030[PDAT]) OR 2019-nCoV[All Fields] OR 2019nCoV[All Fields] OR COVID-19[All Fields] OR SARS-CoV-2[All Fields])

AND (“impact*”[TIAB] OR “effect*”[TIAB])

AND (“policy”[TIAB] OR “policies”[TIAB] OR “order*”[TIAB] OR “mandate*”[TIAB])

AND (“countries”[TIAB] OR “country”[TIAB] OR “state”[TIAB] OR “provinc*”[TIAB] OR “county”[TIAB] OR “parish”[TIAB] OR “region*”[TIAB] OR “city”[TIAB] OR “cities”[TIAB] OR “continent*”[TIAB] “Asia*”[TIAB] OR “Europe*”[TIAB] OR “Africa*”[TIAB] OR “America*”[TIAB] OR “Australia”[TIAB] OR “Antarctica”[TIAB] OR “Afghanistan”[TIAB] OR “Aland Islands”[TIAB] OR “Åland Islands”[TIAB] OR “Albania”[TIAB] OR “Algeria”[TIAB] OR “American Samoa”[TIAB] OR “Andorra”[TIAB] OR “Angola”[TIAB] OR “Anguilla”[TIAB] OR “Antarctica”[TIAB] OR “Antigua”[TIAB] OR “Argentina”[TIAB] OR “Armenia”[TIAB] OR “Aruba”[TIAB] OR “Australia”[TIAB] OR “Austria”[TIAB] OR “Azerbaijan”[TIAB] OR “Bahamas”[TIAB] OR “Bahrain”[TIAB] OR “Bangladesh”[TIAB] OR “Barbados”[TIAB] OR “Barbuda”[TIAB] OR “Belarus”[TIAB] OR “Belgium”[TIAB] OR “Belize”[TIAB] OR “Benin”[TIAB] OR “Bermuda”[TIAB] OR “Bhutan”[TIAB] OR “Bolivia”[TIAB] OR “Bonaire”[TIAB] OR “Bosnia”[TIAB] OR “Botswana”[TIAB] OR “Bouvet Island”[TIAB] OR “Brazil”[TIAB] OR “British Indian Ocean Territory”[TIAB] OR “Brunei”[TIAB] OR “Bulgaria”[TIAB] OR “Burkina Faso”[TIAB] OR “Burundi”[TIAB] OR “Cabo Verde”[TIAB] OR “Cambodia”[TIAB] OR “Cameroon”[TIAB] OR “Canada”[TIAB] OR “Cayman Islands”[TIAB] OR “Central African Republic”[TIAB] OR “Chad”[TIAB] OR “Chile”[TIAB] OR “China”[TIAB] OR “Christmas Island”[TIAB] OR “Cocos Islands”[TIAB] OR “Colombia”[TIAB] OR “Comoros”[TIAB] OR “Congo”[TIAB] OR “Congo”[TIAB] OR “Cook Islands”[TIAB] OR “Costa Rica”[TIAB] OR “Côte d’Ivoire”[TIAB] OR “Croatia”[TIAB] OR “Cuba”[TIAB] OR “Curaçao”[TIAB] OR “Cyprus”[TIAB] OR “Czechia”[TIAB] OR “Denmark”[TIAB] OR “Djibouti”[TIAB] OR “Dominica”[TIAB] OR “Dominican Republic”[TIAB] OR “Ecuador”[TIAB] OR “Egypt”[TIAB] OR “El Salvador”[TIAB] OR “Equatorial Guinea”[TIAB] OR “Eritrea”[TIAB] OR “Estonia”[TIAB] OR “Eswatini”[TIAB] OR “Ethiopia”[TIAB] OR “Falkland Islands”[TIAB] OR “Faroe Islands”[TIAB] OR “Fiji”[TIAB] OR “Finland”[TIAB] OR “France”[TIAB] OR “French Guiana”[TIAB] OR “French Polynesia”[TIAB] OR “French Southern Territories”[TIAB] OR “Futuna”[TIAB] OR “Gabon”[TIAB] OR “Gambia”[TIAB] OR “Georgia”[TIAB] OR “Germany”[TIAB] OR “Ghana”[TIAB] OR “Gibraltar”[TIAB] OR “Greece”[TIAB] OR “Greenland”[TIAB] OR “Grenada”[TIAB] OR “Grenadines”[TIAB] OR “Guadeloupe”[TIAB] OR “Guam”[TIAB] OR “Guatemala”[TIAB] OR “Guernsey”[TIAB] OR “Guinea”[TIAB] OR “Guinea-Bissau”[TIAB] OR “Guyana”[TIAB] OR “Haiti”[TIAB] OR “Heard Island”[TIAB] OR “Herzegovina”[TIAB] OR “Holy See”[TIAB] OR “Honduras”[TIAB] OR “Hong Kong”[TIAB] OR “Hungary”[TIAB] OR “Iceland”[TIAB] OR “India”[TIAB] OR “Indonesia”[TIAB] OR “Iran”[TIAB] OR “Iraq”[TIAB] OR “Ireland”[TIAB] OR “Isle of Man”[TIAB] OR “Israel”[TIAB] OR “Italy”[TIAB] OR “Jamaica”[TIAB] OR “Jan Mayen Islands”[TIAB] OR “Japan”[TIAB] OR “Jersey”[TIAB] OR “Jordan”[TIAB] OR “Kazakhstan”[TIAB] OR “Keeling Islands”[TIAB] OR “Kenya”[TIAB] OR “Kiribati”[TIAB] OR “Korea”[TIAB] OR “Korea”[TIAB] OR “Kuwait”[TIAB] OR “Kyrgyzstan”[TIAB] OR “Lao People’s Democratic Republic”[TIAB] OR “Laos”[TIAB] OR “Latvia”[TIAB] OR “Lebanon”[TIAB] OR “Lesotho”[TIAB] OR “Liberia”[TIAB] OR “Libya”[TIAB] OR “Liechtenstein”[TIAB] OR “Lithuania”[TIAB] OR “Luxembourg”[TIAB] OR “Macao”[TIAB] OR “Madagascar”[TIAB] OR “Malawi”[TIAB] OR “Malaysia”[TIAB] OR “Maldives”[TIAB] OR “Mali”[TIAB] OR “Malta”[TIAB] OR “Malvinas”[TIAB] OR “Marshall Islands”[TIAB] OR “Martinique”[TIAB] OR “Mauritania”[TIAB] OR “Mauritius”[TIAB] OR “Mayotte”[TIAB] OR “McDonald Islands”[TIAB] OR “Mexico”[TIAB] OR “Micronesia”[TIAB] OR “Moldova”[TIAB] OR “Monaco”[TIAB] OR “Mongolia”[TIAB] OR “Montenegro”[TIAB] OR “Montserrat”[TIAB] OR “Morocco”[TIAB] OR “Mozambique”[TIAB] OR “Myanmar”[TIAB] OR “Namibia”[TIAB] OR “Nauru”[TIAB] OR “Nepal”[TIAB] OR “Netherlands”[TIAB] OR “Nevis”[TIAB] OR “New Caledonia”[TIAB] OR “New Zealand”[TIAB] OR “Nicaragua”[TIAB] OR “Niger”[TIAB] OR “Nigeria”[TIAB] OR “Niue”[TIAB] OR “Norfolk Island”[TIAB] OR “North Macedonia”[TIAB] OR “Northern Mariana Islands”[TIAB] OR “Norway”[TIAB] OR “Oman”[TIAB] OR “Pakistan”[TIAB] OR “Palau”[TIAB] OR “Panama”[TIAB] OR “Papua New Guinea”[TIAB] OR “Paraguay”[TIAB] OR “Peru”[TIAB] OR “Philippines”[TIAB] OR “Pitcairn”[TIAB] OR “Poland”[TIAB] OR “Portugal”[TIAB] OR “Principe”[TIAB] OR “Puerto Rico”[TIAB] OR “Qatar”[TIAB] OR “Réunion”[TIAB] OR “Romania”[TIAB] OR “Russian Federation”[TIAB] OR “Rwanda”[TIAB] OR “Saba”[TIAB] OR “Saint Barthélemy”[TIAB] OR “Saint Helena”[TIAB] OR “Saint Kitts”[TIAB] OR “Saint Lucia”[TIAB] OR “Saint Martin”[TIAB] OR “Saint Pierre and Miquelon”[TIAB] OR “Saint Vincent”[TIAB] OR “Samoa”[TIAB] OR “San Marino”[TIAB] OR “Sao Tome”[TIAB] OR “Sark”[TIAB] OR “Saudi Arabia”[TIAB] OR “Senegal”[TIAB] OR “Serbia”[TIAB] OR “Seychelles”[TIAB] OR “Sierra Leone”[TIAB] OR “Singapore”[TIAB] OR “Sint Eustatius”[TIAB] OR “Sint Maarten”[TIAB] OR “Slovakia”[TIAB] OR “Slovenia”[TIAB] OR “Solomon Islands”[TIAB] OR “Somalia”[TIAB] OR “South Africa”[TIAB] OR “South Georgia”[TIAB] OR “South Sandwich Islands”[TIAB] OR “South Sudan”[TIAB] OR “Spain”[TIAB] OR “Sri Lanka”[TIAB] OR “State of Palestine”[TIAB] OR “Sudan”[TIAB] OR “Suriname”[TIAB] OR “Svalbard”[TIAB] OR “Sweden”[TIAB] OR “Switzerland”[TIAB] OR “Syria”[TIAB] OR “Syrian Arab Republic”[TIAB] OR “Tajikistan”[TIAB] OR “Thailand”[TIAB] OR “Timor-Leste”[TIAB] OR “Tobago”[TIAB] OR “Togo”[TIAB] OR “Tokelau”[TIAB] OR “Tonga”[TIAB] OR “Trinidad”[TIAB] OR “Tunisia”[TIAB] OR “Turkey”[TIAB] OR “Turkmenistan”[TIAB] OR “Turks and Caicos”[TIAB] OR “Tuvalu”[TIAB] OR “Uganda”[TIAB] OR “UK”[TIAB] OR “Ukraine”[TIAB] OR “United Arab Emirates”[TIAB] OR “United Kingdom”[TIAB] OR “United Republic of Tanzania”[TIAB] OR “United States Minor Outlying Islands”[TIAB] OR “United States of America”[TIAB] OR “Uruguay”[TIAB] OR “USA”[TIAB] OR “Uzbekistan”[TIAB] OR “Vanuatu”[TIAB] OR “Venezuela”[TIAB] OR “Viet Nam”[TIAB] OR “Vietnam”[TIAB] OR “Virgin Islands”[TIAB] OR “Virgin Islands”[TIAB] OR “Wallis”[TIAB] OR “Western Sahara”[TIAB] OR “Yemen”[TIAB] OR “Zambia”[TIAB] OR “Zimbabwe”[TIAB] OR “Alabama”[TIAB] OR “Alaska”[TIAB] OR “Arizona”[TIAB] OR “Arkansas”[TIAB] OR “California”[TIAB] OR “Colorado”[TIAB] OR “Connecticut”[TIAB] OR “Delaware”[TIAB] OR “Florida”[TIAB] OR “Georgia”[TIAB] OR “Hawaii”[TIAB] OR “Idaho”[TIAB] OR “Illinois”[TIAB] OR “Indiana”[TIAB] OR “Iowa”[TIAB] OR “Kansas”[TIAB] OR “Kentucky”[TIAB] OR “Louisiana”[TIAB] OR “Maine”[TIAB] OR “Maryland”[TIAB] OR “Massachusetts”[TIAB] OR “Michigan”[TIAB] OR “Minnesota”[TIAB] OR “Mississippi”[TIAB] OR “Missouri”[TIAB] OR “Montana”[TIAB] OR “Nebraska”[TIAB] OR “Nevada”[TIAB] OR “New Hampshire”[TIAB] OR “New Jersey”[TIAB] OR “New Mexico”[TIAB] OR “New York”[TIAB] OR “North Carolina”[TIAB] OR “North Dakota”[TIAB] OR “Ohio”[TIAB] OR “Oklahoma”[TIAB] OR “Oregon”[TIAB] OR “Pennsylvania”[TIAB] OR “Rhode Island”[TIAB] OR “South Carolina”[TIAB] OR “South Dakota”[TIAB] OR “Tennessee”[TIAB] OR “Texas”[TIAB] OR “Utah”[TIAB] OR “Vermont”[TIAB] OR “Virginia”[TIAB] OR “Washington”[TIAB] OR “West Virginia”[TIAB] OR “Wisconsin”[TIAB] OR “Wyoming”[TIAB] OR “Ontario”[TIAB] OR “Quebec”[TIAB] OR “Nova Scotia”[TIAB] OR “New Brunswick”[TIAB] OR “Manitoba”[TIAB] OR “British Columbia”[TIAB] OR “Prince Edward Island”[TIAB] OR “Saskatchewan”[TIAB] OR “Alberta”[TIAB] OR “Newfoundland”[TIAB] OR “Labrador”[TIAB])

## Appendix 3: Article review flow diagram

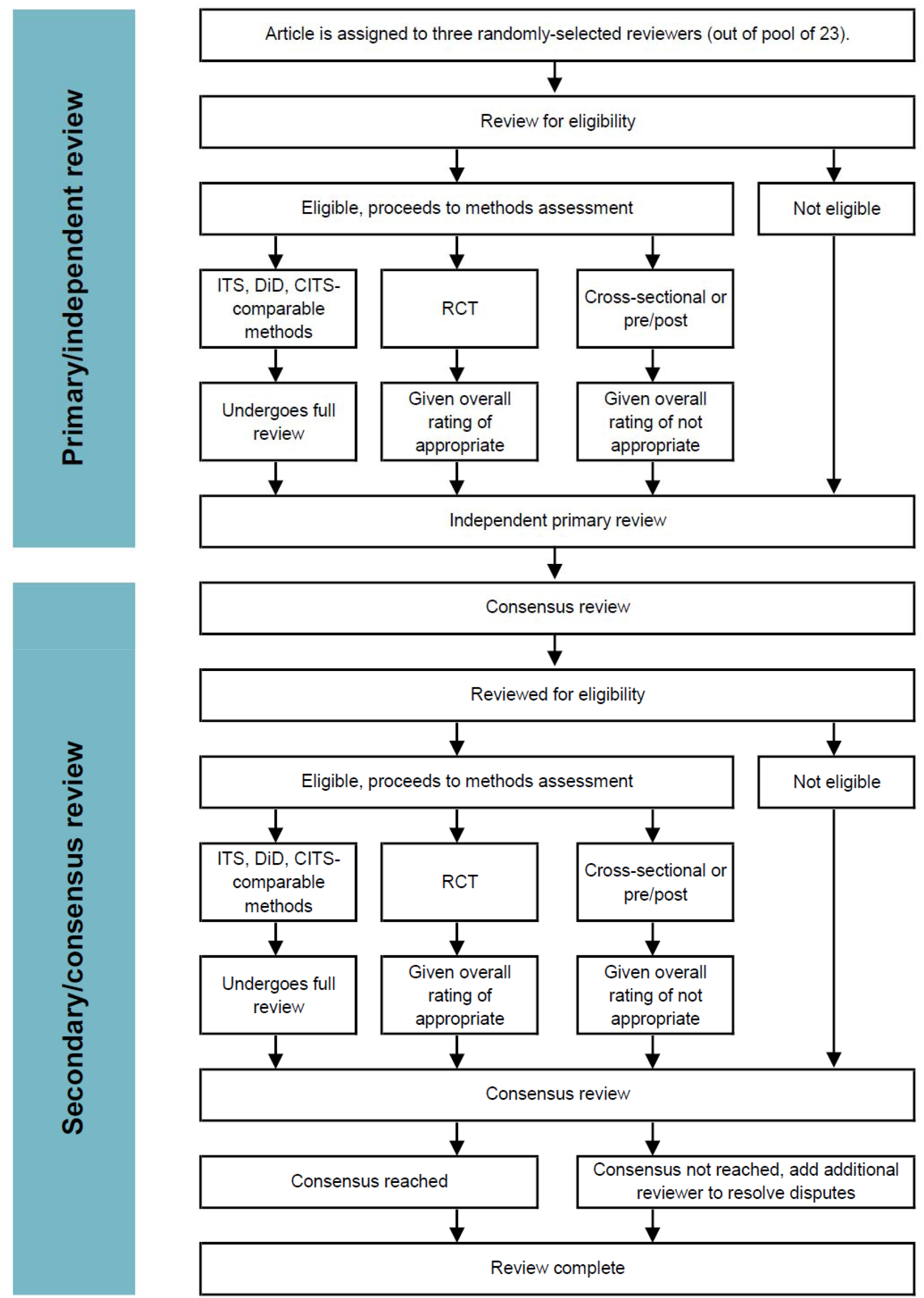

